# Antibody responses to SARS-CoV-2 in patients of novel coronavirus disease 2019

**DOI:** 10.1101/2020.03.02.20030189

**Authors:** Juanjuan Zhao, Quan Yuan, Haiyan Wang, Wei Liu, Xuejiao Liao, Yingying Su, Xin Wang, Jing Yuan, Tingdong Li, Jinxiu Li, Shen Qian, Congming Hong, Fuxiang Wang, Yingxia Liu, Zhaoqin Wang, Qing He, Zhiyong Li, Bin He, Tianying Zhang, Shengxiang Ge, Lei Liu, Jun Zhang, Ningshao Xia, Zheng Zhang

## Abstract

**Background:** The novel coronavirus SARS-CoV-2 is a newly emerging virus. The antibody response in infected patient remains largely unknown, and the clinical values of antibody testing have not been fully demonstrated.

**Methods:** A total of 173 patients with confirmed SARS-CoV-2 infection were enrolled. Their serial plasma samples (n = 535) collected during the hospitalization period were tested for total antibodies (Ab), IgM and IgG against SARS-CoV-2 using immunoassays. The dynamics of antibodies with the progress and severity of disease was analyzed.

**Results:** Among 173 patients, the seroconversion rate for Ab, IgM and IgG was 93.1% (161/173), 82.7% (143/173) and 64.7% (112/173), respectively. Twelve patients who had not seroconverted were those only blood samples at the early stage of illness were collected. The seroconversion sequentially appeared for Ab, IgM and then IgG, with a median time of 11, 12 and 14 days, respectively. The presence of antibodies was < 40% among patients in the first 7 days of illness, and then rapidly increased to 100.0%, 94.3% and 79.8% for Ab, IgM and IgG respectively since day 15 after onset. In contrast, the positive rate of RNA decreased from 66.7% (58/87) in samples collected before day 7 to 45.5% (25/55) during days 15 to 39. Combining RNA and antibody detections significantly improved the sensitivity of pathogenic diagnosis for COVID-19 patients (p < 0.001), even in early phase of 1-week since onset (p = 0.007). Moreover, a higher titer of Ab was independently associated with a worse clinical classification (p = 0.006).

**Conclusions:** The antibody detection offers vital clinical information during the course of SARS-CoV-2 infection. The findings provide strong empirical support for the routine application of serological testing in the diagnosis and management of COVID-19 patients.

## Introduction

Since early December of 2019 and up to February 24, 2020, over 79, 000 cases of coronavirus disease 2019 (COVID-19) caused by novel coronavirus (SARS-CoV-2) infection, with over 2, 600 death cases infection have been reported in 26 countries with a majority of occurrences in China.^1^ The World Health Organization declared a public health emergency of international concern on January 30, 2020. Public health authorities around the world react promptly to prevent further spread of the virus. According to recent reports, most of COVID-19 patients have an incubation period of 3 to 7 days.^2^ Fever, cough and fatigue are the most common symptoms, whereas nasal congestion, runny and diarrhea are only noted in a small part of the patients.^3^ Severe cases might rapidly progress to acute respiratory distress syndrome (ARDS), septic shock and difficult-to-tackle metabolic acidosis and bleeding and coagulation dysfunction.^4^ It should be noted that some of COVID-19 patients only had mild atypical symptoms initially, even for severe and critical cases.^5^ The chest computed tomography features of COVID-19 patients were characterized by the ground-glass opacity and bilateral patchy shadowing.^6^ For laboratory test, it was reported that over 80% of patients had lymphopenia, and most of patients had elevated C-reactive protein.^7^ However, the above-mentioned clinical and laboratory characteristics are not easily distinguishable from pneumonia induced by infection with other common respiratory tract pathogens such as influenza virus, streptococcus pneumoniae and mycoplasma pneumoniae.

The timely and accurate diagnosis of the SARS-CoV-2 infection is the cornerstone of the efforts to provide appropriated treatment for patients, to limit further spread of the virus and ultimately to eliminate the virus from human society. Currently, viral RNA detection by several polymerase chain reaction (PCR) based technics is almost the only way to confirm the diagnosis of SARS-CoV-2 infection in practice. On the other hand, RNA testing based on throat or nasopharyngeal swabs brought out negligible false-negative risk.^8^ The reported positive rate varied for different swab specimens in COVID-19 patients. ^3,9^ Many cases that were strongly epidemiologically linked to SARS-CoV-2 exposure and with typical lung radiological findings remained RNA negative in their upper respiratory tract samples. There are four potential reasons: 1) the viral loads in upper respiratory tract samples are much lower than that in lower respiratory tract samples in COVID-19 patients;^9^ 2) the releasing viral loads of patients in different stage of infection varies with a wide range;^10^ 3) the collection of high-quality swab specimen requires skillful health-workers; and 4) PCR reagents from different sources have high variance. Consequently, these problems lead to a noteworthy delay of early diagnosis and following management and propose serious challenge to providing timely life support treatment and preventive quarantine.

For many known pathogenic viruses, it has been a routine practice to make a diagnosis of acute infection according to the serological findings for a long time. Comparing to PCR, serological testing is advantageous with faster turn-around time, high-throughput and less workload. However, the clinical value of antibodies largely depends on the understanding of host antibody responses during infection. Given that SARS-CoV-2 is a newly emerging virus, the antibody response in COVID-19 patients remains largely unknown. To mitigate this knowledge gap, and to provide scientific analysis on the benefit of antibody testing when used in combination with the current RNA testing, this study investigates the dynamics of total antibody (Ab), IgM and IgG antibody against SARS-CoV-2 in serial blood samples collected from 173 confirmed COVID-19 patients and provides discussion on the clinical value of antibody testing.

## Methods

### Patients

A confirmed COVID-19 case and the clinical classification was defined based on the New Coronavirus Pneumonia Prevention and Control Program (4th edition) published by the National Health Commission of China. This study enrolls a total of 173 cases of COVID-19, where all patients were admitted to the Shenzhen Third People’s Hospital between Jan 11 and Feb 9, 2020, and were willing to donate their blood samples. All enrolled cases were confirmed to be infected with SARS-CoV-2 by use of real-time RT-PCR (rRT-PCR) on samples from the respiratory tract. For all enrolled patients, the date of illness onset, clinical classification, RNA testing results during the hospitalization period, and the personal demographic information were obtained from the clinical records. This study was reviewed and approved by the Medical Ethical Committee of Shenzhen Third People’s Hospital (approval number 2020-0018). Written informed consent was obtained from each enrolled patient.

### Antibody measurement

The total antibody (Ab), IgM antibody and IgG antibody against SARS-CoV-2 in plasma samples were tested using enzyme linked immunosorbent assay (ELISA) kits supplied by Beijing Wantai Biological Pharmacy Enterprise Co., Ltd., China, according to the manufacturer’s instructions. Briefly, the ELISA for total antibodies detection was developed based on double-antigens sandwich immunoassay (Ab-ELISA), using mammalian cell expressed recombinant antigens contained the receptor binding domain (RBD) of the spike protein of SARS-CoV-2 as the immobilized and HRP-conjugated antigen. The IgM μ-chain capture method (IgM-ELISA) was used to detect the IgM antibodies, using the same HRP-conjugate RBD antigen as the Ab-ELISA. The IgG antibodies were tested using indirect ELISA kit (IgG-ELISA) based on a recombinant nucleoprotein antigen. The specificity of the assays for Ab, IgM and IgG was determined as 99.1% (211/213), 98.6% (210/213) and 99.0% (195/197) by testing of samples collected from healthy individuals before the outbreak of SARS-CoV-2.

### Statistical analysis

For continue variables description, mean with standard deviation was used for normal distribution data and median with interquartile range (IQR) was used for non-normal distribution data. Cumulative seroconversion rates were calculated by Kaplan-Meier method. The association between antibody level and severity of illness were estimated by generalized estimating equations (GEE) model with logit link function. All statistical analysis was conducted by SAS 9.4 (SAS Institute, Cary, NC, USA).

## Results

### The characterization of patients

Among totally 368 cases of COVID-19 patients admitted in the hospital (before Feb 9, 2020), 173 patients of them (47%) were enrolled in the study (Table 1). All patients had acute respiratory infection syndromes and/or abnormalities in chest CT images accompanied by detectable SARS-CoV-2 RNA in respiratory sample since illness onset for at least one time. The median age of the studied patients was 48 years (IQR, 35-61 years) and 51.4% were females. There were 126 patients had clear epidemiological travel/residence history in Wuhan (116) and other cities of Hubei province (10), respectively (Table 1). Among them, 32 (18.5%) were in critical illness condition with ARDS or oxygen saturation < 93% who required mechanical ventilation either invasively or non-invasively, and the remaining 141 (81.5%) had mild to moderate syndromes were in non-critical condition. By February 19, a total of 62 patients (35.8%,54 were in non-critical group and 8 were in critical group) were recovered and discharged from hospital and 2 (1.1%, both were in critical group) patients died with underlying chronic disease.

**Table 1.**
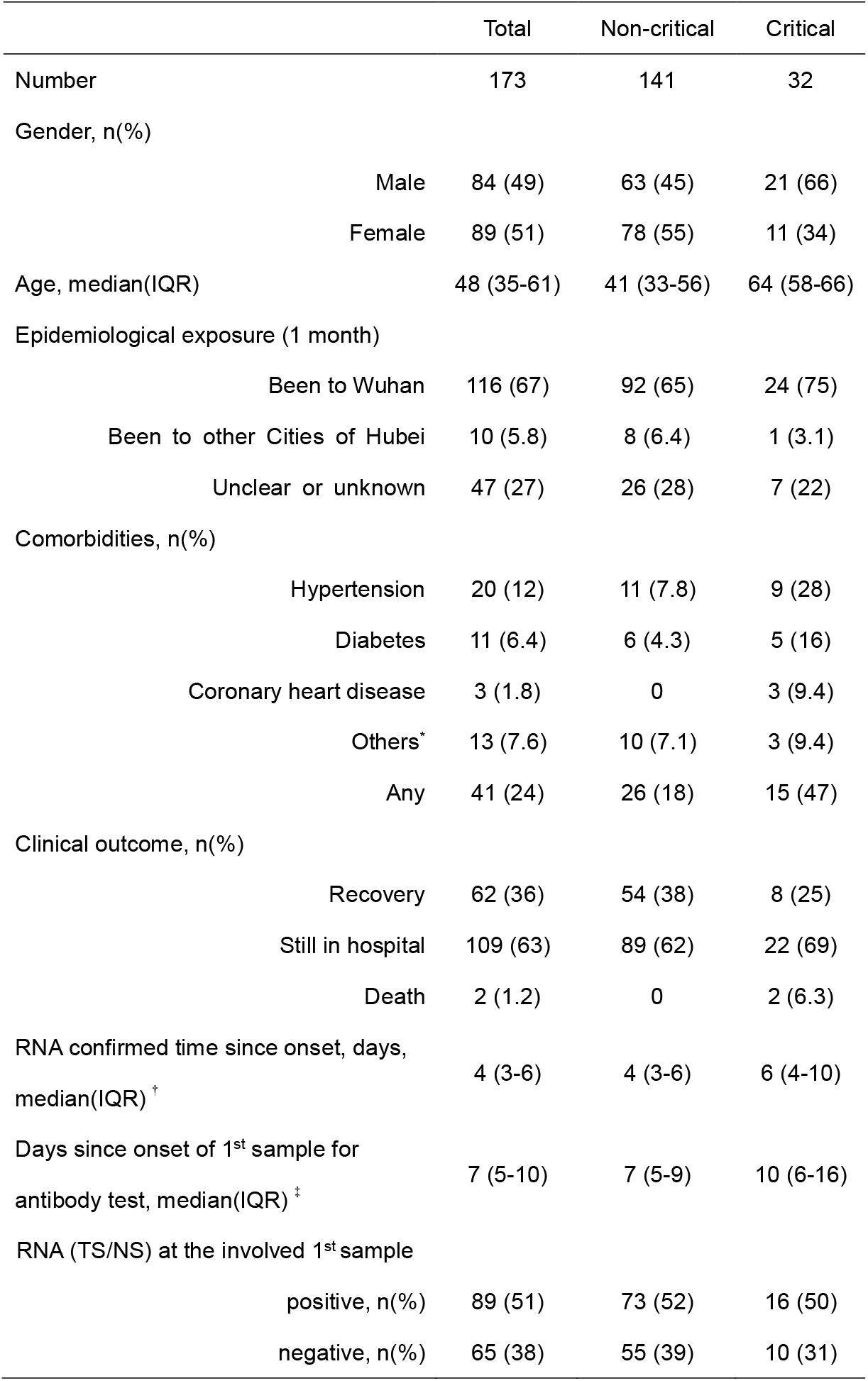
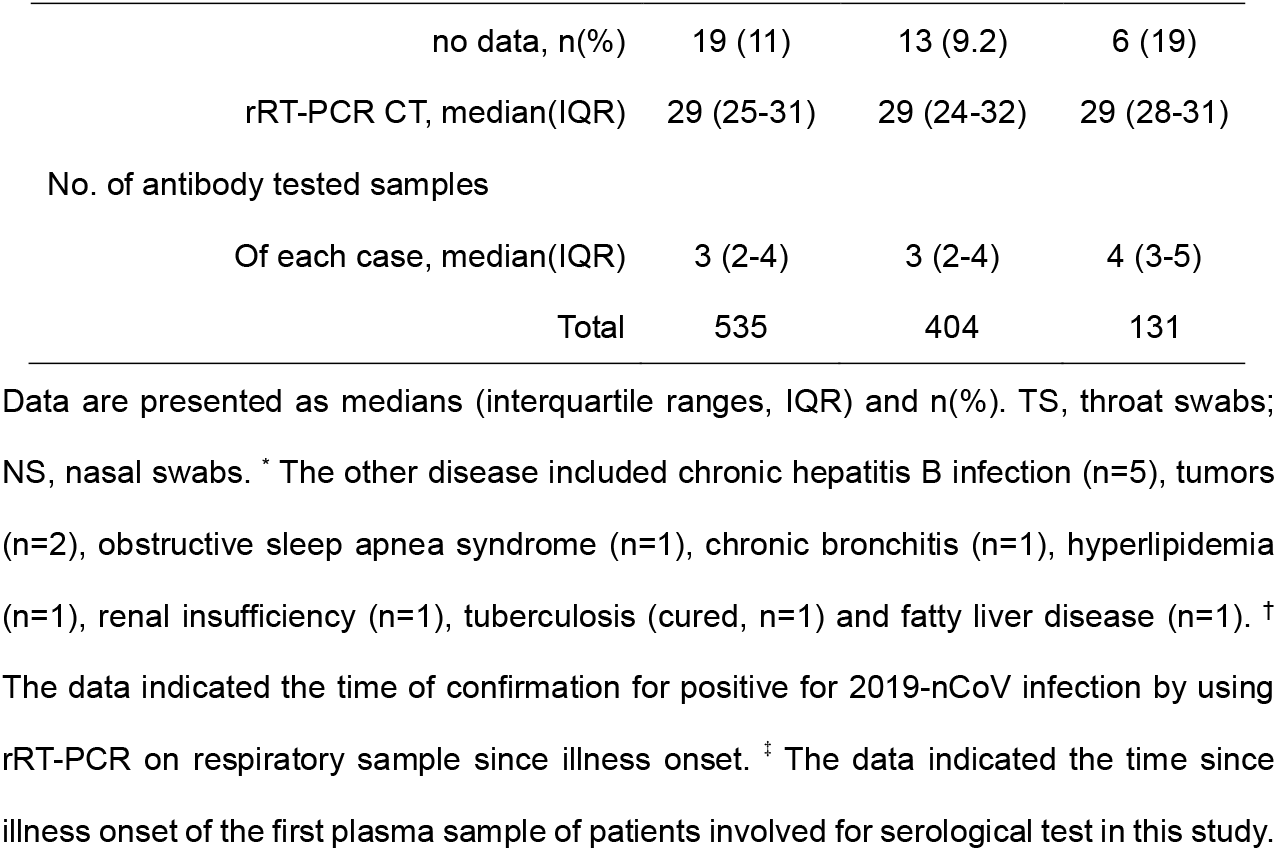
Demographics and clinical characteristics of patients and sample cohort with COVID-19 in this study.

### Seroconversion of antibodies against SARS-CoV-2 in COVID-19 patients

A total of 535 plasma samples collected during the hospitalization period of the 173 patients were tested for antibodies against SARS-CoV-2. The seroconversion rate for Ab, IgM and IgG was 93.1% (161/173), 82.7% (143/173) and 64.7% (112/173), respectively (Table 1). Twelve patients who remained seronegative for Ab testing possibly due to that their samples involved were all collected at the early stage of illness (10 earlier than day 10, the other two on day 11 and 13 after onset). The cumulative seroconversion curve showed that the rate for Ab and IgM reached 100% around 1 month of illness day. The seroconversion was sequential appeared for Ab, IgM and then IgG (Figure 1A). The median time to Ab, IgM and IgG seroconversion was 11, 12 and 14 days, separately. One of two patients tested on the onset day was seropositive. Overall, the seroconversion of Ab was significantly quicker than that of IgM (p = 0.012) and IgG (p < 0.001), that possibly attributed to the double-antigen sandwich form of the assay used which usually show much higher sensitivity than capture assay (IgM) and indirect assay (IgG). Moreover, all isotypes of antibodies against viral antigen, including IgM, IgA and IgG, can be detected by double-sandwich based assay, which may also contribute to the superior performance of Ab test. In comparisons of seroconversion rates of antibodies between critical and non-critical patients, none of the three markers showed significant difference (Figure 1B, 1C and ID, p > 0.05).

**Figure 1.**
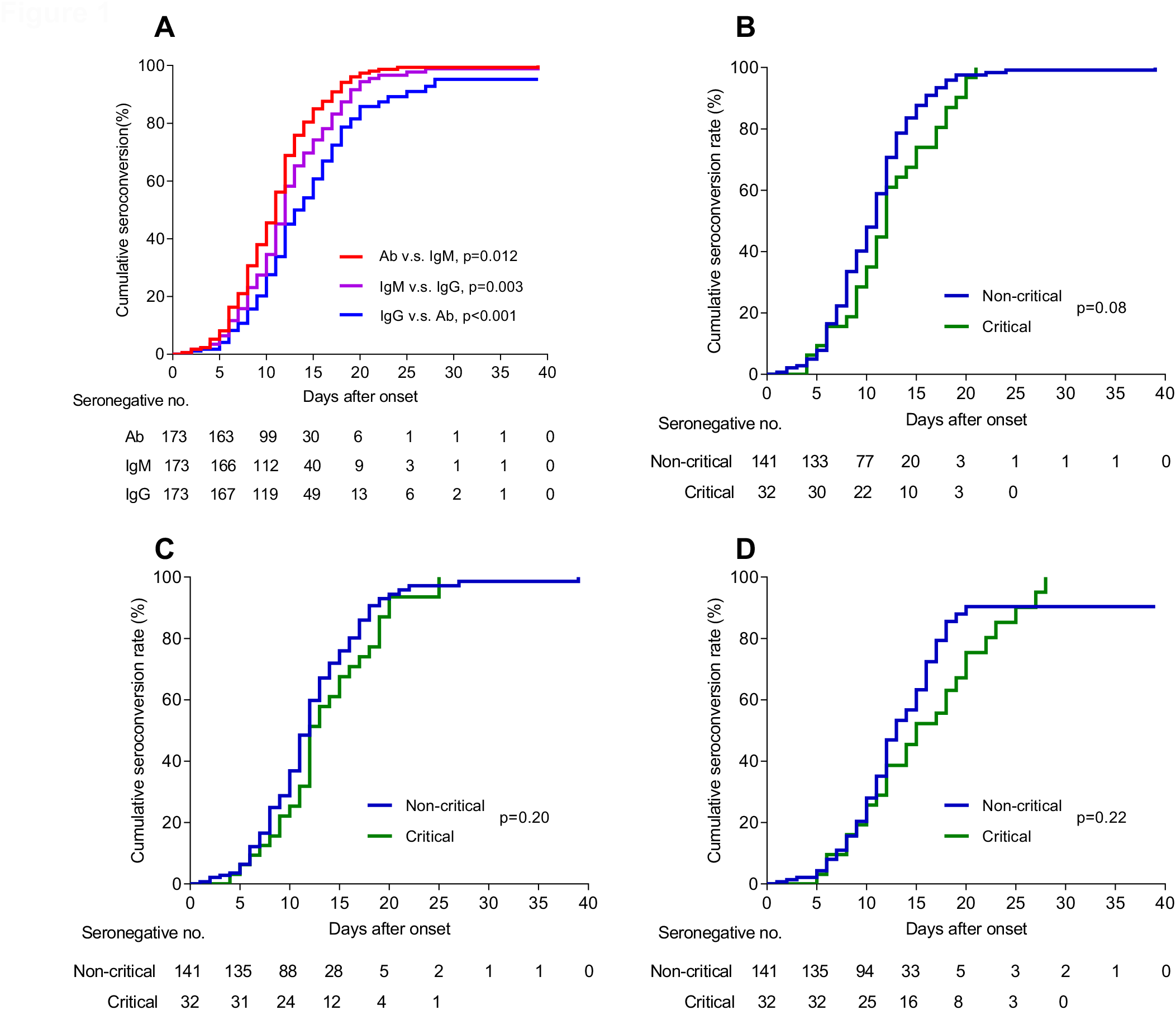
Cumulative incidence of seroconversion of antibodies against SARS-CoV-2 among COVID-19 patients during the acute phase since illness onset. (A) Cumulative incidence of seroconversion of Ab, IgM and IgG among 173 patients of this study. Kaplan–Meier curves for time to seroconversion of Ab (B), IgM (C) and IgG (D) for comparison of patients in critical condition or not. P values were determined by Log-Rank test to compare different markers.

### The diagnosis value of antibody assays for patients in different time after onset

We analyzed the detectability of RNA test and antibody assays according to the time course since illness onset in the cohort. As the results shown in Figure 2 and Table 2, in the early phase of illness within 7-day since onset, the RNA test had the highest sensitivity of 66.7%, whereas the antibody assays only presented a positive rate of 38.3%. However, the sensitivity of Ab overtook that of RNA test since day 8 after onset and reached over 90% across day 12 after onset (Figure 2). In samples from patients during day 8-14 after onset, the sensitivities of Ab (89.6%), IgM (73.3%) and IgG (54.1%) were all higher than that of RNA test (54.0%). Among samples from patients in later phase (day 15-39 since onset), the sensitivities of Ab, IgM and IgG were 100.0%, 94.3% and 79.8%, respectively. In contrast, RNA was only detectable in 45.5% of samples of day 15-39. Further analyses demonstrated that in patients with undetectable RNA in their respiratory tract samples collected during day 1-3, day 4-7, day 8-14 and day 15-39 since onset, there were 28.6% (2/7), 53.6% (15/28), 98.2% (56/57) and 100% (30/30) had detectable antibody in total Ab assay, respectively (Table 3). Whatever, combined use of the tests of RNA and Ab improved markedly the sensitivities of pathogenic-diagnosis for COVID-19 patients in different phases (Table 2, Ab+RNA).

**Table 2.** Performance of different detections in samples at different time since onset of patients.

| Days |  | RNA |  | Ab |  | IgM |  | IgG |  | RNA+Ab |  |
| --- | --- | --- | --- | --- | --- | --- | --- | --- | --- | --- | --- |
| after | n |  | Sensitivity |  | Sensitivity |  | Sensitivity |  | Sensitivity |  | Sensitivity |
| onset |  | n(+) | (%, 95%CI) | n(+) | (%, 95%CI) | n(+) | (%, 95%CI) | n(+) | (%, 95%CI) | n(+) | (%, 95%CI) |
| Total | 173 | 112<sup>\$</sup> | 67.1<br>(59.4, 74.1) | 161 | 93.1<br>(88.2, 96.4) | 143 | 82.7<br>(76.2, 88) | 112 | 64.7<br>(57.1, 71.8) | 172 | 99.4<br>(96.8, 100.0) |
| 1-7 | 94 | 58<sup>\$</sup> | 66.7<br>(55.7, 76.4) | 36 | 38.3<br>(28.5, 48.9) | 27 | 28.7<br>(19.9, 39.0) | 18 | 19.1<br>(11.8, 28.6) | 74 | 78.7<br>(69.1, 86.5) |
| 8-14 | 135 | 67<sup>\$</sup> | 54.0<br>(44.8, 63.0) | 121 | 89.6<br>(83.2, 94.2) | 99 | 73.3<br>(65.0, 80.6) | 73 | 54.1<br>(45.3, 62.7) | 131 | 97.0<br>(92.6, 99.2) |
| 15-39 | 90 | 25<sup>\$</sup> | 45.5<br>(32.0, 59.5) | 90 | 100.0<br>(96.0, 100.0) | 83<sup>*</sup> | 94.3<br>(87.2, 98.1) | 71<sup>#</sup> | 79.8<br>(69.9, 87.6) | 90 | 100.0<br>(96.0, 100.0) |

\* Two patients missed IgM tests due to inadequate plasma samples. # One patient missed IgG tests due to inadequate plasma samples. \$ There were 7, 11 and 35 patients had not been performed RNA testing during the 1-7 onset day, 8-14 onset day and 15-39 onset day, respectively.

**Table 3.** Serological presence of antibodies against SARS-CoV-2 in patients with undetectable viral RNA at different time since onset of disease.

| Days after<br>onset | No. of patients<br>with undetectable RNA* | Detectable antibody in plasma, n (%) |  |  |
| --- | --- | --- | --- | --- |
|  |  | Ab | IgM | IgG |
| 1-3 | 7 | 2 (28.6) | 2 (28.6) | 2 (28.6) |
| 4-7 | 28 | 15 (53.6) | 12 (42.9) | 8 (28.6) |
| 8-14 | 57 | 56 (98.2) | 45 (78.9) | 40 (70.2) |
| 15-39 | 30 | 30 (100) | 28 (93.3) | 22 (73.3) |

\* RNA was tested using throat/nasal swab sample.

**Figure 2.**
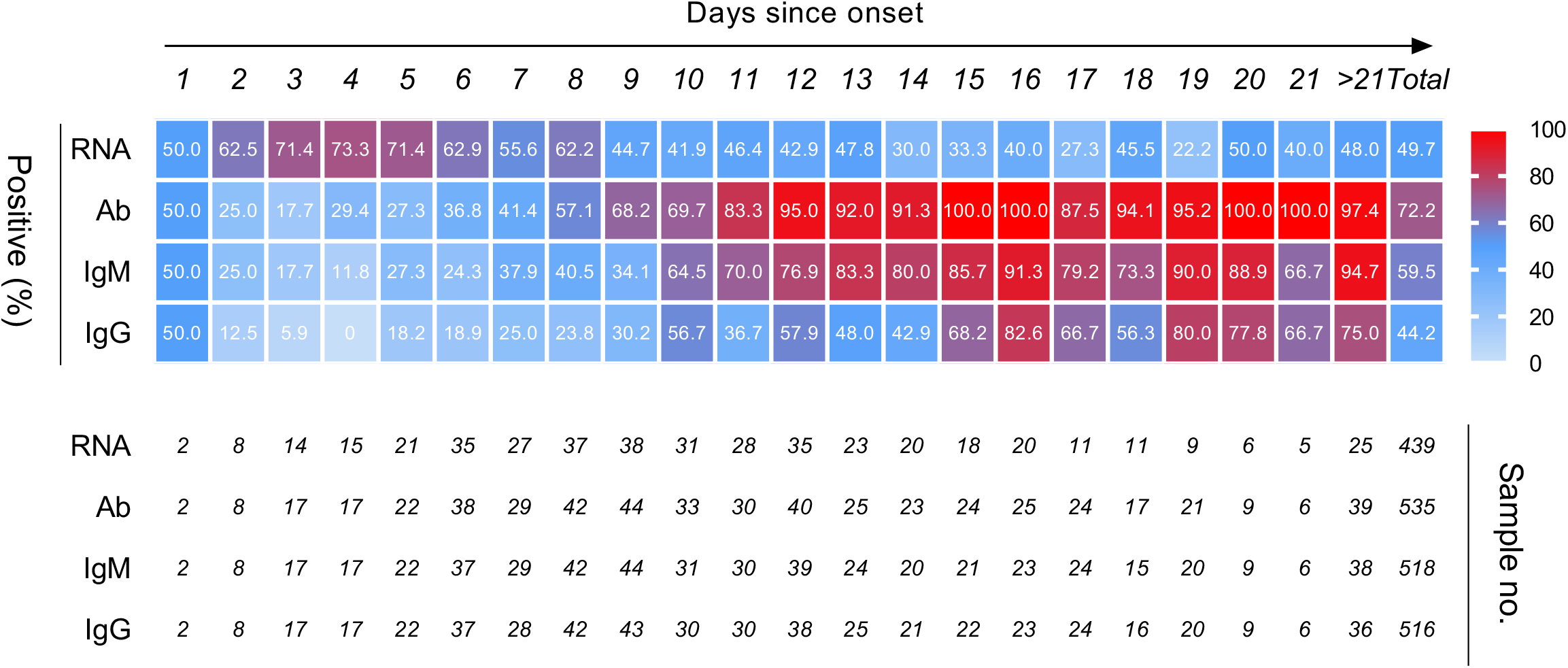
Profiling of sensitivity performances of RNA, Ab, IgM and IgG in time series since illness onset. A heat-map of detection of SARS-CoV-2 infection according to the time (days) since onset by a single RNA or antibody test.

### The dynamics of antibody levels with the progress and severity of disease

To investigate the dynamics of antibody level according disease course, the antibody levels were expressed using the relative binding signals compared to the cutoff value of each assay (S/CO). The longitudinal changes of antibody and RNA in 9 representative patients, including 6 in non-critical group (Figure 3A) and 3 in critical group (Figure 3B), were presented in Figure 3. The first positive time point of RNA tests appeared earlier than that of Ab in 7 of 9 patients, except for the case 185 (Ab was detectable 2 days earlier than RNA) and the case 111 (at the same day). It should be noted that the risings of antibodies were not always accompanied by RNA clearance, particularly in the 3 critical patients. This finding suggested that antibodies may not be sufficient to clear the virus. In the pooled analyses on all involved patients, the average antibody levels showed a marked increase since about 1-week after onset and continuously elevated during the next 2 weeks (Figure 4A). Further analyses suggested that there was no significant difference on the average S/CO value of Ab tests between critical and non-critical patients before day 12 after onset (Figure 4B). However, critical patients showed significantly higher Ab S/CO values than non-critical cases in about 2-week after onset (p = 0.02) and this association was not significant in either IgM or IgG tests (data not shown). For further exploration, we determined the relative Ab titer of these samples (expressed as relative optical density, rOD) by serial dilution measurements of each sample. The quantitative data of Ab titers also revealed a significant difference (p = 0.004) between patients in critical and non-critical groups (Figure 4C). Multivariate longitudinal GEE analyses suggested that age (β = 0.139, p < 0.001), gender (β = 1.415, p = 0.006) and Ab titer (β = 0.336, p = 0.006) were the independent factors strongly associated with the clinical classification based on the severity (Table 4).

**Table 4.**
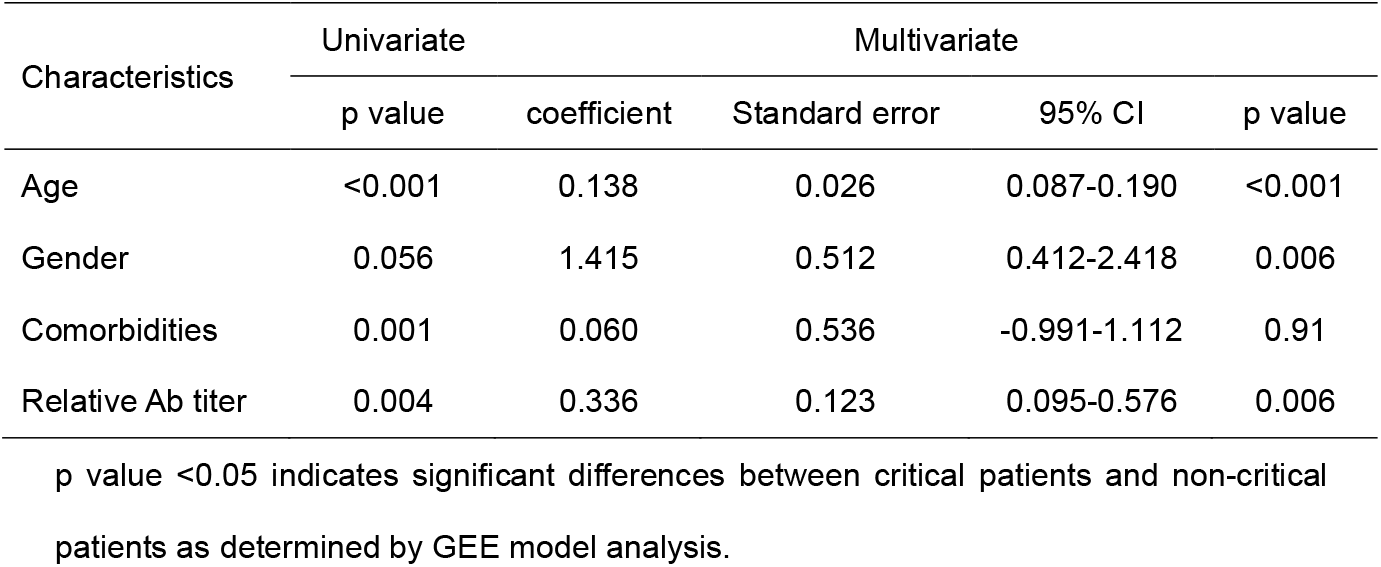
Univariate and multivariate analysis of factors associated with patients in critical condition.

**Figure 3.**
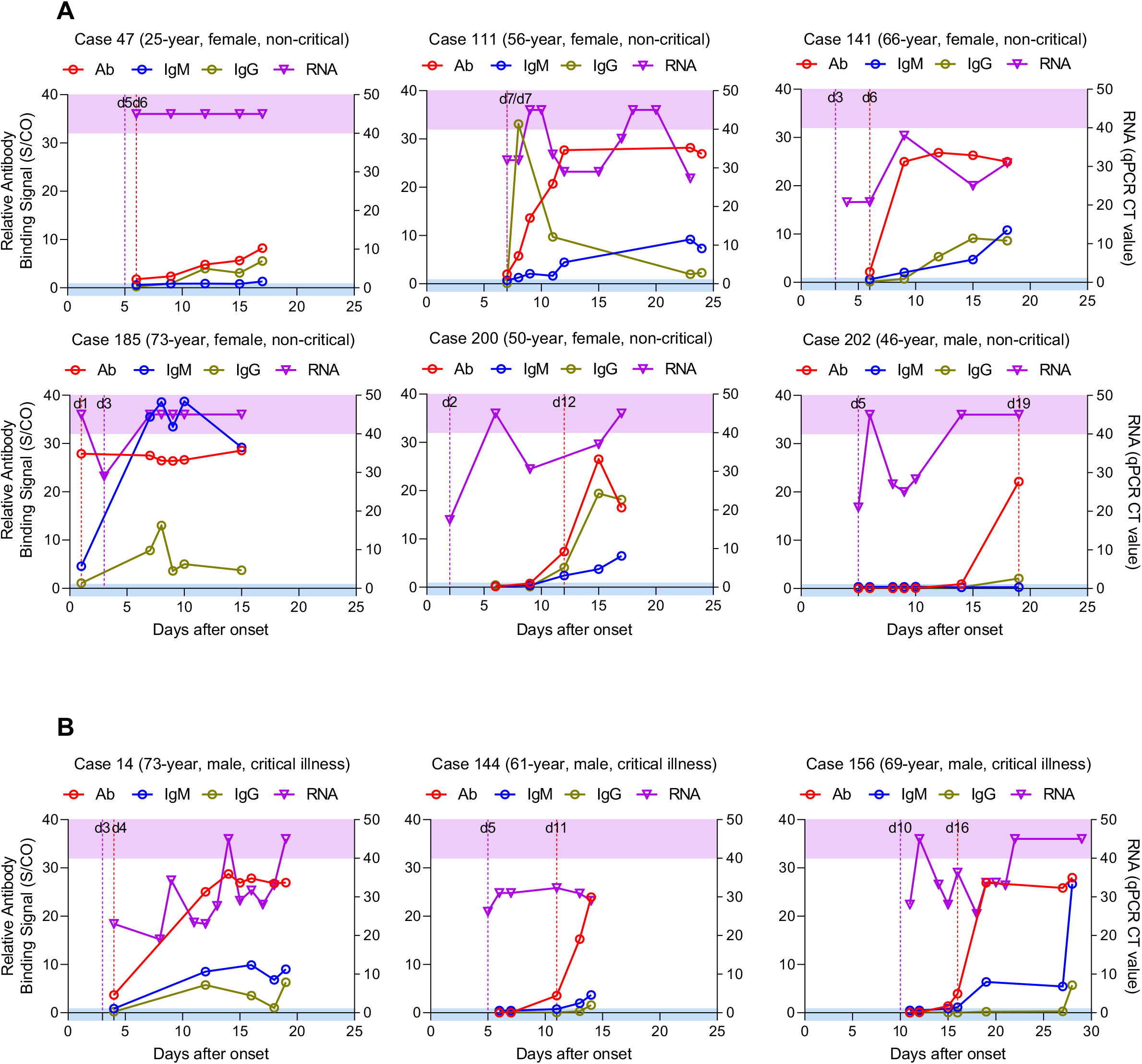
Dynamic profiling of viral RNA and antibodies in representative COVID-19 patients since onset of disease. The changes of the levels of RNA in in upper respiratory specimens (nasal and/or throat swabs) and antibodies (total Ab, IgM and IgG) in plasma of 9 patients were presented. Among these cases, 6 were in normal to moderate illness condition (A) and 3 were in critical condition (B). The cutoff values for antibody tests were S/CO=1 (plotted to left-Y axis) and was CT=40 for RNA detection (plotted to right-Y axis). RNA negative samples are denoted with a CT of 45. The blue area indicated the antibody seronegative zone, whereas the purple area indicated undetectable RNA zone. Meanwhile, a purple broken line was used to indicate the first time point with detectable RNA and a red broken line was used to indicate the first antibody seroconversion (total Ab) time point.

**Figure 4.**
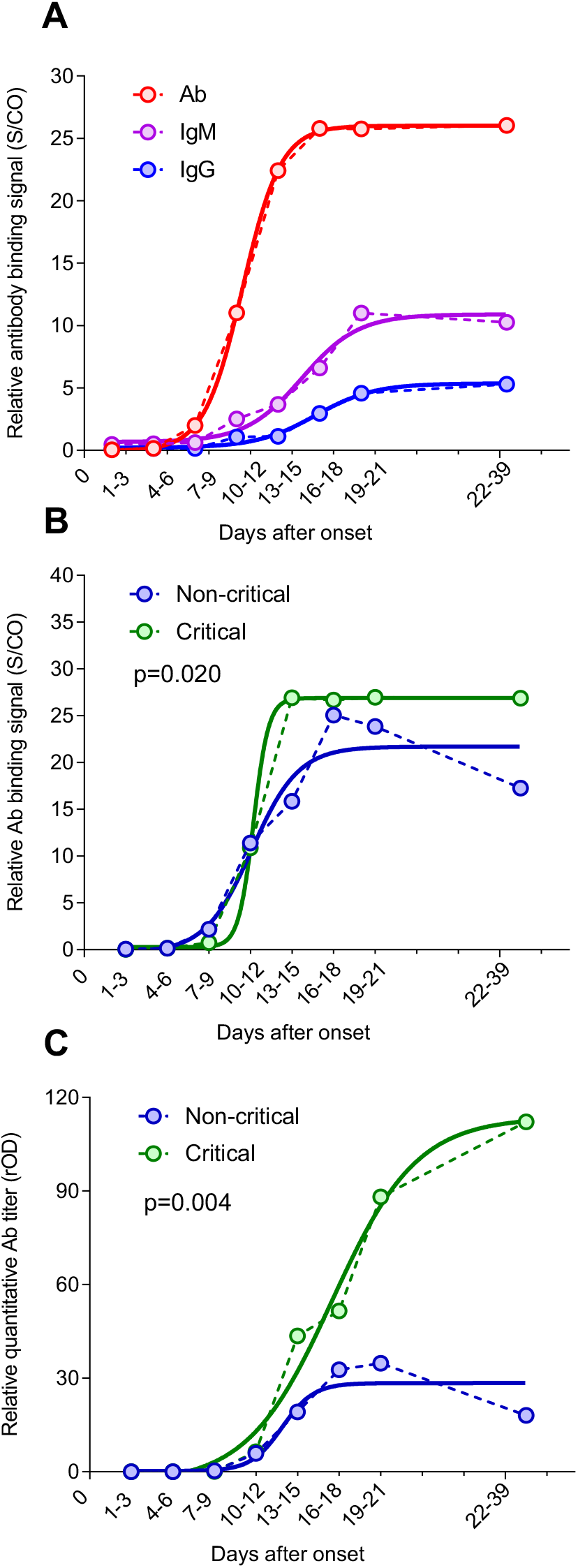
The average levels of antibodies against SARS-CoV-2 among COVID-19 patients since illness onset. (A) Comparison of the average S/CO value of in between total Ab, IgG and IgM. Comparison of the average S/CO value (B) and relative quantitative titer (C) of Ab test between critical and non-critical patients. The medians of antibody detection value (S/CO for tests of Ab, IgM and IgG, for panel A and B) and of total Ab titer (rOD, for panel C) of samples at the same time point since onset was used to plot the graph. Patient’s samples collected from day 1-3, 4-6, 7-9, 10-12, 13-15, 16-18, 19-21, 22-39 since illness onset were pooled for analysis. Four parameter logistic (4PL) fitting curves were used to show the rising trend of antibodies (total Ab, IgG, IgM).

## Discussion

The present data demonstrated that typical antibody responses to acute viral infection are wildly induced in COVID-19 patients. All patients were seroconverted, except for 12 patients with only samples collected at the early stage of illness (before 13 day of onset). To be expected, the total antibody was first detected, followed by IgM and IgG. The seroconversion rate and the antibody levels increased rapidly during the first two weeks, the cumulative seropositive rate reached 50% on the 11th day and 100% on the 39th day. The seroconversion time of total antibody, IgM and IgG antibodies appeared consequently (p < 0.05) with a median seroconversion day of 11, 12 and 14, respectively. Due to the lack of blood samples collected from patients in the later stage of illness, how long the antibodies could last remained unknown. Our results demonstrated an excellent sensitivity of Ab test in detections of patient’s samples after 1-week since onset. Notably, even in the early stages of the illness within 7-day, some patients with negative nucleic acid findings could be screened out through antibody testing. Combining RNA test and antibody test significantly raised the sensitivity for detecting patients (p < 0.001). Above findings indicate that the antibody detection be an important supplement to RNA detection during the illness course.

Up to date, the confirm diagnosis of SARS-CoV-2 infection entirely depend on the viral nucleic acid testing. Even though with high analytical sensitivity, the real-world performance of PCR based RNA testing is unsatisfied. Many suspected patients had to be tested for several days with multiple samples before confirm diagnosis were made, and during the waiting time they might have not enough priority to receive relevant treatments and quarantine managements.^3^ These problems make the timely diagnosis of SARS-CoV-2 infection one of the bottlenecks for adapting relevant actions to limit the damage of current outbreak. Our study provided robust evidences that: 1) the acute antibody response in SARS-CoV-2 infection patient is very similar to many other acute viral infections; 2) the serological testing can be a powerful approach in achieving timely diagnosis of SARS-CoV-2 infection; and 3) the total antibody is more sensitive than IgM and IgG for detecting SARS-CoV-2 infection.

Thus, the antibody testing might play vital roles in the following settings: 1) for the suspected patient under the initial visit or clinically diagnosed patients has not been confirmed by RNA testing, the positive result of antibody increases the confidence to make a COVID-19 diagnosis; 2) for healthy close contact who is in the quarantine period, he/she should be deemed as a probable carriers if antibody positive, then the RNA should be tested more frequently and the close contacts of him/her should be observed; 3) for the RNA confirmed patient, seropositive indicates that the specific immune response had been induced. Besides, epidemiological studies could be conducted using immunoassays. Additional, it can play an important role in searching potential animal hosts for SARS-CoV-2 using Ab-ELISA because the double antigen sandwich method makes it free from species restriction. It has been less than three months since the SARS-CoV-2 virus first invaded human society, and the prevalence of antibody against SARS-CoV-2 is nearly zero. Therefore, at least during the current outbreak which is likely to continue to May or June 2020, individuals who are seropositive could be a probably preceding infector of SARS-CoV-2. During this short period, therefore, the total antibody could be considered as a recent infection marker similar as IgM antibody. Our data showed that the sensitivity of total antibody testing is higher than IgM or IgG. Therefore, the total antibody detection should be given high priority to be implemented in current clinical and public health practice. If, unfortunately, SARS-CoV-2 become a common respiratory transmission pathogen lasting in human society, such as influenza viruses or low-pathogenicity types of coronaviruses, rather than be completely eradicated as its relative SARS-CoV-1 virus, the serological diagnosis of acute SARS-CoV-2 infection will more depend on the detection of IgM antibody in post-epidemic areas, such as Wuhan, China, in the subsequent epidemic seasons. The total antibody and IgG antibody could be used to understand the epidemiology of SARS-CoV-2 infection and to assist in determining the level of humoral immune response in patients. Even then, the total antibody will be a more sensitive marker for sentinel monitoring of imported cases in naive community.

In addition to the diagnosis value of Ab test, our study revealed a strong positive correlation between clinical severity and antibody titer since 2-week after illness onset, for the first time in COVID-19 patients. The results suggested that a high titer of total antibodies against the virus may be considered as a risk factor of critical illness, independently from older age, male gender and comorbidities (Table 4). Although it was still unclear how the causal relation between hormonal response and illness severity, the results raise a possible usage of the high antibody titer as a surrogate marker for worse clinical prognosis. Furthermore, it might be an evidence for the possibility of antibody-dependent disease enhancement effects, which was commonly found in SARS-CoV-1 patients.^11-13xs^ Whatsoever, our finding suggested that the clinical meanings of the level of antibody against SARS-CoV-2 during the acute phase of infection warrant further study.

It should be noted that there were some limitations of this study. First, for most of RNA tests of the patients were based on upper respiratory tract specimens, the positive rate may be higher in detection using lower respiratory tract specimens, such as bronchoalveolar lavage fluid, deep tracheal aspirates, and induced sputum may yield higher sensitivity for RNA tests. Second, we cannot evaluate the persistence of antibodies because samples were collected during the acute illness course of patients. Third, although it had shown good specificity in healthy people, the cross-reactivity among the different coronaviruses cannot be accurately assessed because we cannot obtain blood samples from SARS-CoV-1 and other coronaviruses infection patients. Future studies are needed to a better understanding of the antibody response profile of SARS-CoV-2 infection.

In conclusion, the findings demonstrate that antibody tests have important diagnosis value in addition to RNA tests. These findings provide strong evidence for the routine application of serological antibody assays in the diagnosis and clinical management of COVID-19 patients.

## Data Availability

We will share individual participant data that underlie the results reported in this article, after deidentification (text, tables, figures and appendices). Also redacted Study Protocol and Statistical Analysis Plan will be shared. The data will be available beginning 6 months after the major findings from the final analysis of the study were published, ending 2 years later. The data will be shared with investigators whose proposed use of the data has been approved by an independent review committee identified for individual participant data meta-analysis. Proposals should be directed to. To gain access, data requestors will need to sign a data access agreement.

## Contributors

ZZ, NS-X, JZ, LL, S-X G and QY had the idea and for designed the study. ZZ, JZ, LL and S-X G had full access to all data in the study and take responsibility for the integrity of the data and the accuracy of the data analysis. JJ-Z, HY-W, XJ-L, XW, JY, JZ-L, QS, FX-W, YX-L, ZQ-W, QH, LL and ZZ had roles in the clinical management, patient recruitment, sample preparation and clinical data collection. JJ-Z, HY-W, WL, XW, TD-L, CM-H, ZY-L, BH and SX-G had roles in the antibody detection experiments, data collection and analysis. QY, YY-S, SX-G, JZ and ZZ had roles in statistical analysis. JJ-Z, QY, YY-S, SX-G, LL, JZ, NS-X and ZZ had roles in data interpretation. QY, YY-S, SX-G, JZ and ZZ wrote the manuscript. TY-Z, NS-X and ZZ contributed to critical revision of the report. All authors reviewed and approved the final version of the manuscript.

## Acknowledgements

We acknowledge the work and contribution of all the health providers from Shenzhen Third People’s Hospital. We sincerely thanked the Shan Qiao, Xue-Rong Jia, Dong Wang and Bao-Liang Jia from Beijing Wantai Biological Pharmacy Company for their helpful technical assistance. This study was supported by Bill & Melinda Gates Foundation.

